# National Patterns of Remote Patient Monitoring Service Availability at US Hospitals

**DOI:** 10.1101/2024.10.14.24315496

**Authors:** Aline F. Pedroso, Zhenqiu Lin, Joseph S. Ross, Rohan Khera

## Abstract

**Background:** Digital remote patient monitoring (RPM) enables longitudinal care outside traditional healthcare settings, especially in the vulnerable period after hospitalizations, with broad coverage of the service by payers. We sought to evaluate patterns of RPM service availability at US hospitals and the characteristics of hospitals and the communities they serve that are associated with the availability of these services.

**Methods:** We used contemporary national data from the American Hospital Association (AHA) Annual Survey to ascertain US hospitals offering RPM services for post-discharge or chronic care. We linked hospitals with their census-based county-level data to define the characteristics of the communities they serve and examined the association of these characteristics with RPM availability. We also conducted an exploratory analysis quantifying the number of patients with key cardiovascular conditions of heart failure (HF) and acute myocardial infarction (AMI) receiving care at hospitals providing RPM services.

**Results:** The study included 5,644 hospitals. Over five years of study, there was a 40.3% increase in the number of hospitals offering RPM services, rising from 1,364 (33.0%) hospitals in 2018 to 1,797 (46.3%) in 2022. However, the availability of RPM services varied across different hospital groups, with smaller, non-teaching, rural, and hospitals serving low-income communities, particularly in the South, being less likely to offer RPM. Hospitals with more than 300 beds had 3.7-fold odds of offering RPM compared with those with less than 100 beds (aOR 3.71, 95% CI 2.90-4.74). Non-teaching hospitals had a 70% lower likelihood of RPM availability (aOR 0.29, 95% CI 0.19–0.44), and rural hospitals had 50% lower odds compared with urban hospitals (aOR 0.49, 95% CI 0.32-0.77).

**Conclusions:** In this national study of US hospitals, there has been a large increase in the availability of RPM services but with large variation among hospitals, with lower availability in hospitals serving low-income and rural communities.

**What is Known:**

- Remote patient monitoring (RPM) allows longitudinal care outside traditional healthcare settings through digital technologies and is increasingly reimbursed by payers.
- RPM services have been shown to support care quality and improve access to care and are now endorsed by international clinical guidelines as a core component of cardiovascular disease management.
- RPM utilization in the United States has grown in recent years, driven in part by the introduction of new billing codes and policy reforms.

**What the Study Adds:**

- This is the first national hospital-level analysis to assess the availability of RPM services and identify hospital and community characteristics associated with their adoption.
- RPM availability has grown significantly, but uptake remains lower among smaller, rural, non-teaching hospitals and those serving disadvantaged populations.
- Despite overall increase in RPM availability, the gap observed between urban and rural hospitals persisted over time, indicating that growth in adoption did not translate into a narrowing of geographic disparities in access.

## INTRODUCTION

Access to emerging digital health services in the US remains highly variable across hospitals, often reflecting broader disparities in healthcare resources, technological infrastructure, and institutional capacity.^1,2^ These differences can directly influence patient outcomes, including hospital readmission rates. Digital technologies that enable the monitoring of patients outside clinical settings, or remote patient monitoring (RPM), present a promising avenue to address the persistent challenge of preventable hospital readmissions, particularly for cardiovascular conditions. However, the extent to which hospitals are adopting RPM services and the factors associated with the availability of these services remain poorly understood.^3,4^

Contemporary RPM services represent payer-reimbursed care wherein patients use in-home or wearable devices to remotely collect digital health information, such as their weight, heart rate, and blood pressure, which are transmitted electronically for review by clinicians.^5,6^ These data can enable clinicians to identify early markers of clinical deterioration and intervene before patients suffer hospitalization.^7,8^ The evaluation of RPM services has demonstrated its role in maintaining the quality of care, facilitating faster access to services, reducing patient travel costs, and minimizing the frequency of clinic visits.^4,9,10^ Their use has also been endorsed by European clinical practice guidelines, which now recommend remote monitoring as a standard component of disease management programs for cardiovascular conditions like HF.^11^ In the US, while there has been a growing emphasis on value-based care, it is unclear whether disparities in RPM adoption exist across hospitals, particularly in relation to the characteristics of the communities they serve.^6,12,13^

In this study, we pursued a national assessment of the availability of RPM services at US hospitals. We examined the association between the characteristics of hospitals and the communities they serve with the availability of RPM services. In an exploratory analysis, we assessed the number of fee-for-service Medicare beneficiaries that would be affected by the availability of RPM services when hospitalized with heart failure (HF) and acute myocardial infarction (AMI).

## METHODS

All data and materials have been made publicly available at the GitHub repository and can be accessed at [https://github.com/CarDS-Yale/rpm_national].

### Data Sources and Study Population

We identified hospitals offering RPM services using data from the American Hospital Association (AHA) Annual Survey and defined their respective county-level population characteristics based on the US census. Additional information about the AHA Annual Survey can be found in **Supplemental Methods**. We used data from the AHA Annual Surveys from 2018 through 2022, representing the most recent 5-year period that collected information on RPM services.^22^ We used the US Cities Database as an up-to-date record of all US cities and towns to link hospitals to their corresponding serving communities’ characteristics, based on the county where the hospital is located. All demographic information is derived from the American Community Survey of the US Census Bureau.^23^

To evaluate the number of Medicare beneficiaries affected by access to RPM service, we quantified the number of Medicare fee-for-service beneficiaries discharged with a principal diagnosis of HF or AMI. We obtained hospitalization counts using publicly reported data on hospital case volumes in the Center of Medicare & Medicaid Services (CMS) Hospital Quality Reporting program. We linked them to AHA data using unique Medicare Provider Identification Numbers.^25^

### RPM service availability

The AHA survey defined RPM as the use of digital technologies to collect medical and other forms of health data from individuals in one location and electronically transmit the information securely to healthcare providers in a different location for assessment and recommendation.^22^ The survey specifically inquired about the availability of RPM in the context of post-discharge care, ongoing care for chronic conditions, and remote monitoring in other clinical scenarios. The primary exposure for the study was the availability of any RPM service to patients at a hospital.

### Covariates

We identified a series of hospital characteristics from the AHA survey. These included hospital bed size, presented as the absolute number of available beds as well as categorized into subsets with <100, 100-300, and >300 beds, US region (Northeast, Midwest, South, and West), and area type (metropolitan, micropolitan, and rural, based on the definitions of the US Census Bureau). According to the US Census Bureau, a metropolitan statistical area is a core area with an urbanized area of 50,000 or more, plus adjacent counties with high social and economic ties. A micropolitan statistical area has a core urban cluster of 10,000 to 49,999, also with adjacent counties linked through commuting and other ties. Rural areas, as defined by the Census Bureau, encompass all population, housing, and territory not included within an urban area. For exploratory analyses, we further classified hospital location by rural-urban commuting area (RUCA) codes for 2010, which classify US census tracts using measures of population density, urbanization, and daily commuting. We subclassified hospitals into metropolitan (RUCA codes, 1-3), micropolitan (RUCA codes, 4-6), small (RUCA codes, 7-9), and rural (RUCA code, 10) based on the RUCA codes corresponding to their Federal Information Processing Standards (FIPS) code. Hospital’s teaching status was defined based on their membership in the Council of Teaching Hospitals of the Association of American Medical Colleges, and hospital ownership as either government or privately owned.

Key demographic and socio-economic characteristics of the communities served by these hospitals included the distribution of age (proportion of individuals 65 years and older), gender (proportion of females), race and ethnicity (proportion of self-declared Black or Hispanic), median household income in US Dollars, education level (proportion of people with less than a high school education), and the proportion of disabled individuals in their respective counties.

### Statistical analysis

We pursued three key analyses. First, we described the availability of RPM services across all hospitals for each study year. We examined the characteristics of hospitals and the communities served by these hospitals with and without the availability of RPM services and assessed the temporal trends in their availability from 2018 to 2022. Categorical features are presented as counts and percentages, with group-level differences compared using the chi-square test for proportions. Continuous features are presented as means and standard deviations or median and interquartile ranges, with group-level differences compared using the non-parametric Kruskal-Wallis test.

Second, we assessed hospital and community-level factors independently associated with the availability of RPM services in a cross-sectional assessment in 2022, the most recent year in the analysis. For this, we employed a mixed-effects logistic regression model. The dependent variable in this model was a hospital’s RPM service availability, with hospital (bed size, US region, area type, teaching status, ownership) and population characteristics (proportion of individuals aged 65 and older, proportion of females, proportion of self-reported Black and Hispanic individuals, proportion of people with less than a high school education, proportion of disabled individuals, median household income) as key independent variables, and the hospital site as a random effect. Continuous county-level characteristics were modeled as a function of a single standard deviation unit of change for each characteristic. We specifically examined the relative availability of RPM services across US hospitals based on location (metropolitan, micropolitan, small town, or rural areas) and calendar year, using analysis of covariance with the RPM availability as the dependent variable, hospital location as an independent variable, and an interaction term for hospital location and year.

We then explored the potential clinical reach of RPM services by quantifying the distribution of HF and AMI patients hospitalized across institutions with and without RPM services from 2018 to 2022. For each year, we aggregated hospital-level discharge counts and calculated the total number and proportion of patients discharged from hospitals offering RPM services to their patients. These proportions were used to evaluate trends in RPM coverage among patients with cardiovascular conditions over time. We used the Cochran-Armitage test for trend to assess whether there was a statistically significant increase in these proportions over this 5-year period.

We used Python version 3.12.3 and Stata 18 for all analyses. The level of significance was set at 0.05. Since the study uses aggregate data across hospitals without any patient-level information, it was outside the purview of the Yale Institutional Review Board.

## RESULTS

### Hospital Characteristics

There were 5,644 acute care hospitals in the AHA Survey included in the study. Among these hospitals, the median bed size was 80 (IQR 28-196), 12.8% were in the Northeast, and 66.6% were in metropolitan areas. A total of 257 (4.6%) were teaching hospitals.

Of these hospitals, 2,198 (38.9%) reported having RPM services available for one or more years during the study period. There was a 40.2% increase in the number of hospitals offering RPM services over the 5 years, increasing from 33.0% in 2018 to 46.3% in 2022 (**Table 1**). Over the study years, RPM availability was higher among certain hospital subgroups. For instance, a higher proportion of hospitals with >300 beds had RPM services available than those with <100 beds (>300 beds 54.5% in 2018 to 72.0% in 2022, vs <100 beds 24.1% to 34.2%), though the availability of RPM services increased broadly between 2018 and 2022 (32.1% increase among hospitals with >300 beds and 41.8% among hospitals with <100 beds). Hospitals in the Northeast had the highest RPM service availability across study years (47.9% in 2018 to 62.8% in 2022) with a relative increase in availability of 31.0%, with those in the South with the lowest availability (32.7% in 2018 and 47.2% in 2022, 45.5% relative increase in RPM availability over the 5 years of study). Similarly, teaching hospitals, those located in urban areas, and privately-owned hospitals had higher RPM service availability than rural, non-teaching, and government-owned hospitals, respectively (**Table 1**, **Figure 1, Table S2, Figures S1 & S2**). **Figure 2** shows the distribution of hospitals offering RPM services across US counties.

**Figure 1.**
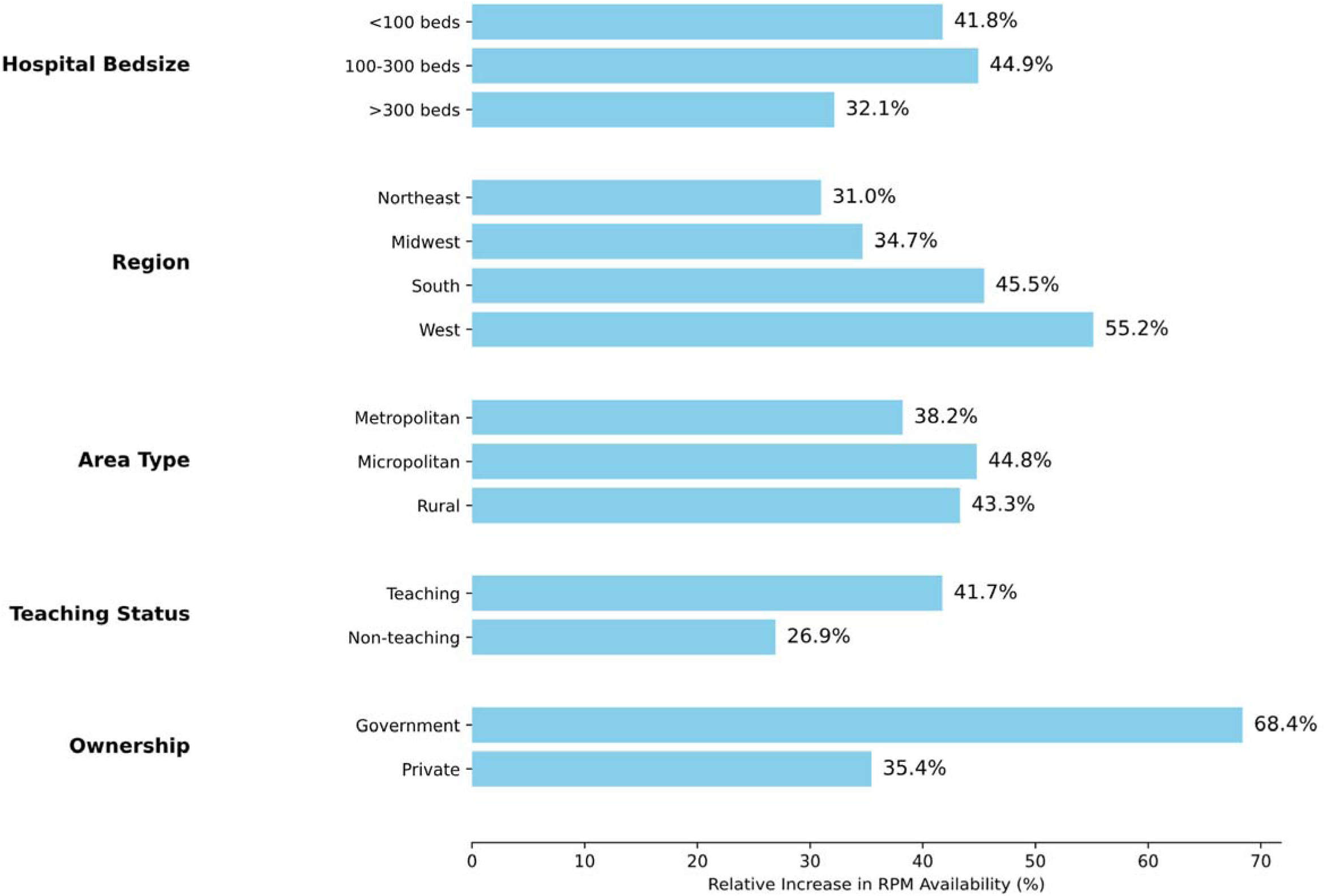
Relative increase in remote patient monitoring (RPM) services across hospital groups between 2018 and 2022.

**Figure 2.**
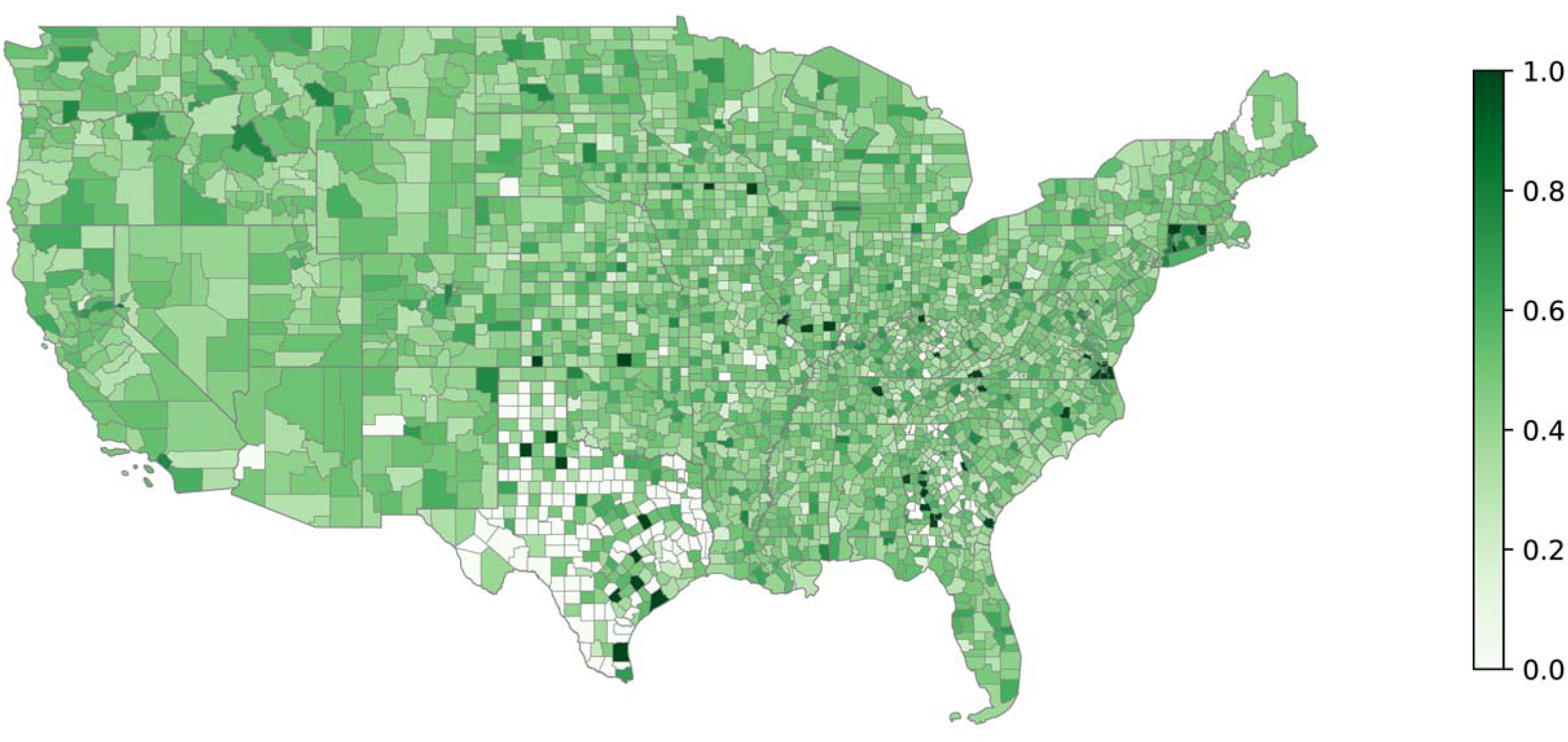
Geographic distribution of ZIP codes with hospitals offering remote patient monitoring (RPM) services across US counties. Color intensity reflects the proportion of ZIP codes with hospitals offering RPM relative to the total ZIP codes in each county.

**Table 1.**
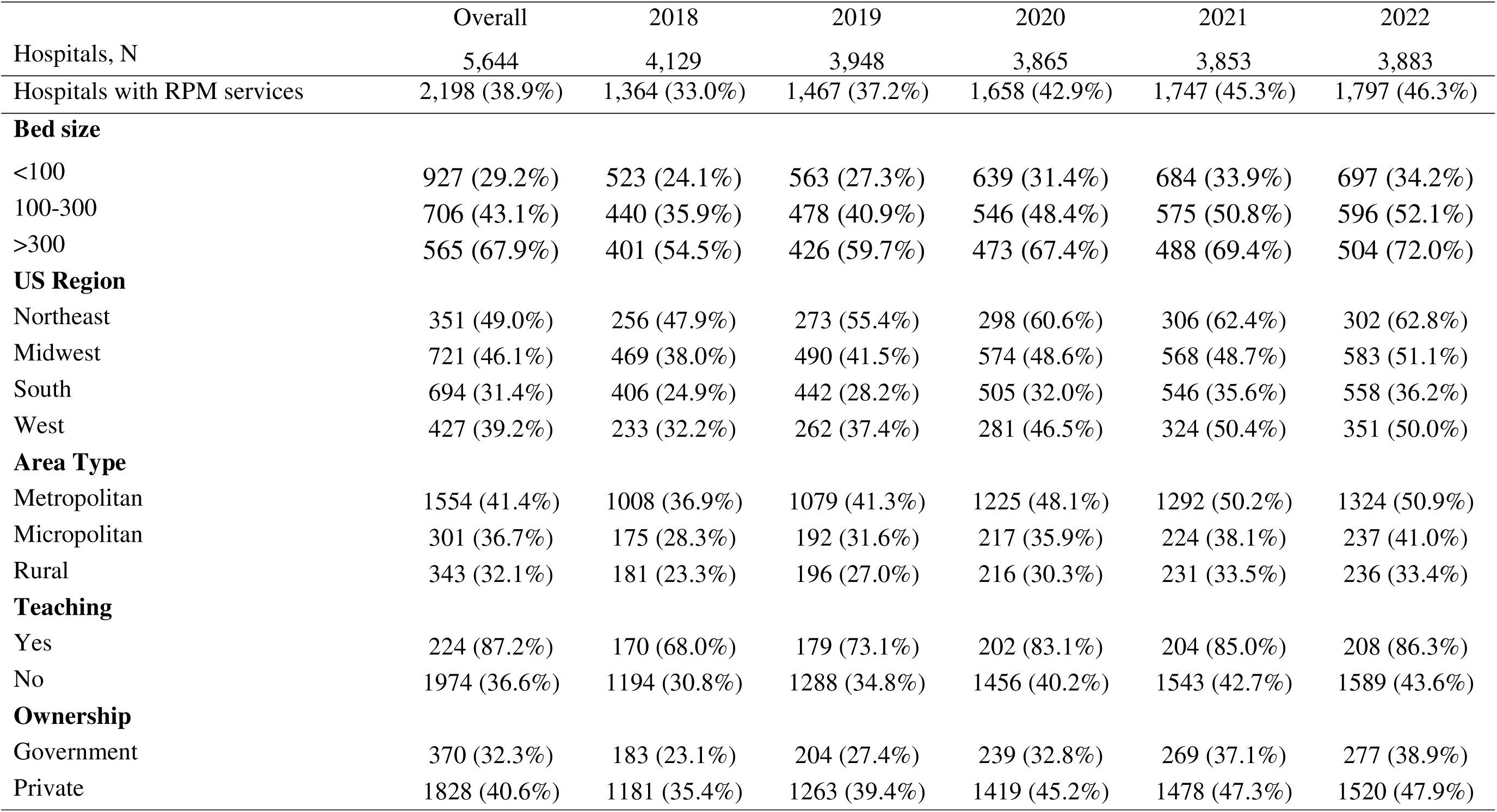
Hospitals with remote patient monitoring (RPM) services between 2018 and 2022. For individual hospital characteristics, the numbers and percentages represent those offering RPM services in the respective hospital subgroups.

The proportion of hospitals offering RPM increased steadily across all RUCA categories from 2018 to 2022, with no evidence that trends differed by RUCA classification. The greatest increase was observed in metropolitan areas (rising from 35% to nearly 50%) with persistently lower adoption in rural hospitals (increasing from 23% to 33%).

### Community Characteristics

A median of 14.3% (IQR 12.6-16.3) of the population in the counties served by these hospitals were 65 years or older, and 51.0% (IQR 50.1-51.9) were female. The proportion of Black and Hispanic individuals in these areas represented a median 16.3% (IQR 5.8-28.8) and 12.5% (IQR 5.9-29.0) of the population, respectively. The proportion of individuals with less than a high school education was a median 11.9% (IQR 8.8-16.2) across counties, and the median annual household income was $60,187.0 (IQR $50,284-$71,673). The median age, gender, and race/ethnicity distributions were similar between neighborhoods with and without hospitals offering RPM services to their patients. However, hospitals offering RPM services were in counties with higher median household incomes and lower percentages of individuals with less than a high school education (**Table 2**).

**Table 2.**
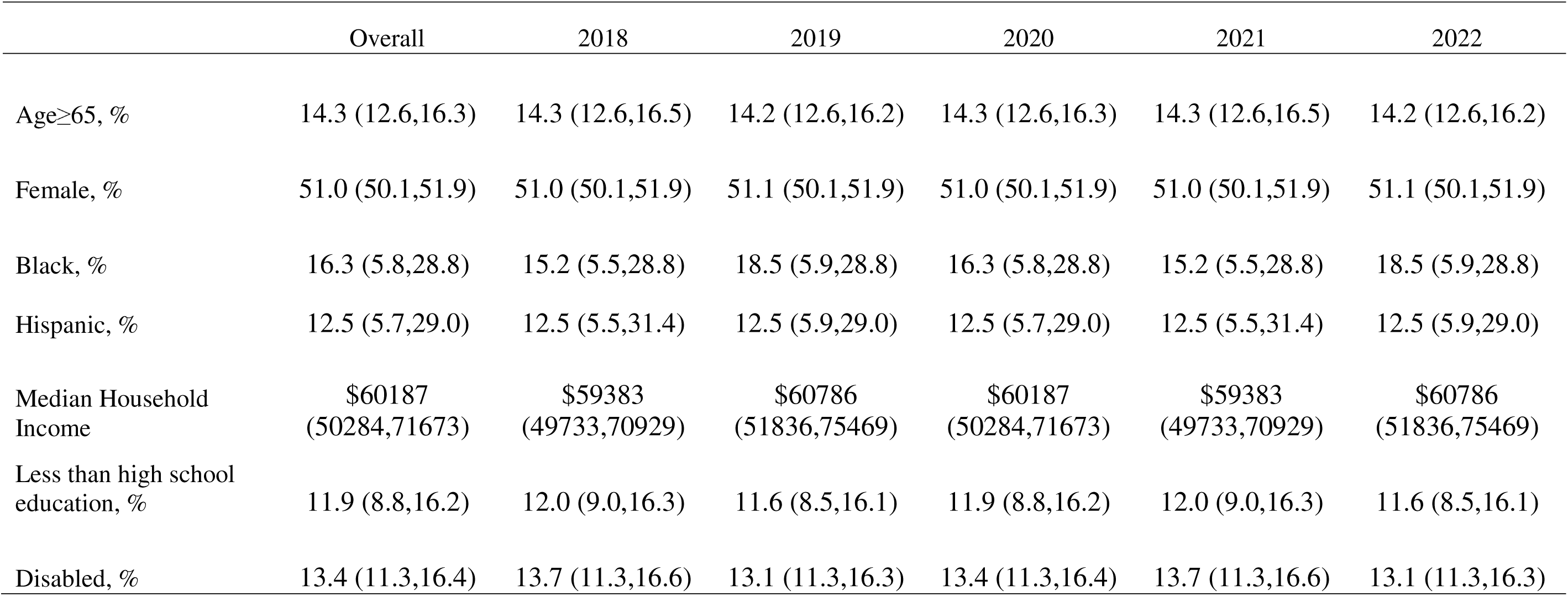
Population characteristics of counties with hospitals with remote patient monitoring services (2018-2022).

### Factors associated with RPM service availability

In adjusted analyses, hospital bed size, teaching status, rural/urban location, and geographic region were significantly associated with the availability of RPM services (**Figure 3, Figures S3 & S4**). Hospitals with more than 300 beds had higher odds of RPM service availability than those with fewer than 100 beds (adjusted OR (aOR) 3.71, 95% CI 2.90-4.74). Non-teaching hospitals had over 70% lower odds of offering RPM services than teaching hospitals (aOR 0.29, 95% CI 0.19-0.44), and rural hospitals had 50% lower odds of RPM availability compared with hospitals in urban metropolitan areas. Additionally, hospitals in the South and West were less likely to have RPM services than those in the Northeast (aOR 0.38, 95% CI 0.29-0.49, and aOR 0.58, 95% CI 0.43-0.78, respectively).

**Figure 3.**
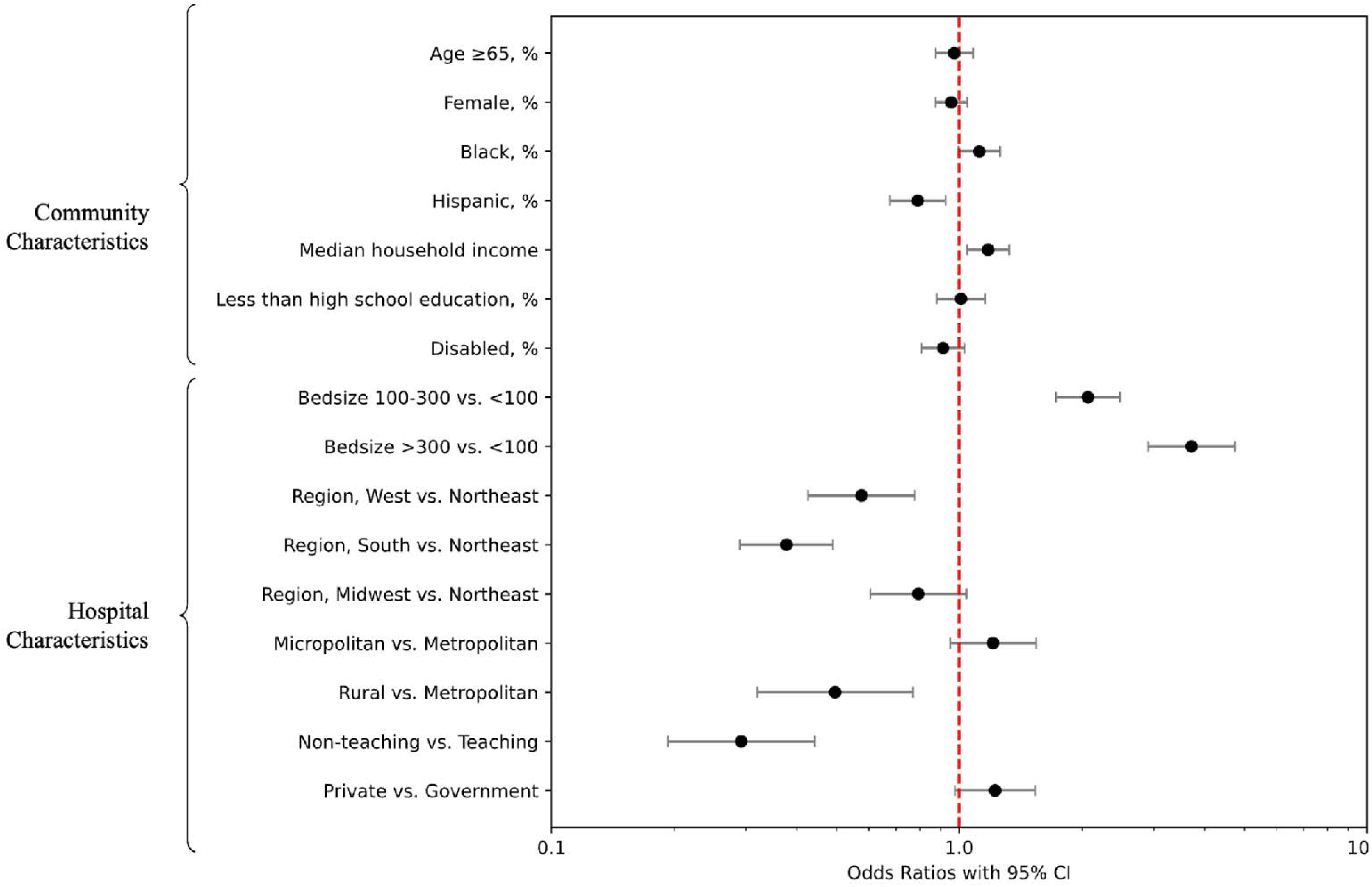
Community and hospital characteristics associated with the availability of remote patient monitoring services.

For characteristics of communities served by hospitals, the proportion of Hispanic individuals and the median income of the community were key features associated with the availability of RPM services at their local hospitals. Each standard deviation increase in the percentage of Hispanic individuals in the community was associated with 20% lower odds of hospitals in that county having RPM services (aOR 0.79, 95% CI 0.68-0.93). Conversely, higher area-level median household income was associated with 18% higher odds of RPM service availability at hospitals in those communities (aOR 1.18, 95% CI 1.05-1.33, per standard deviation unit change). Other hospital and population characteristics were not significantly associated with RPM availability (**Table S1**).

### Burden of Cardiovascular Patients at Hospitals with and without RPM Services

The proportion of hospitalizations for HF and AMI for Medicare fee-for-service beneficiaries at hospitals with RPM services increased over time. In 2018, 52.9% and 55.1% of HF and AMI admissions, respectively, were at hospitals that offered RPM, and increased to 71.4% and 72.4% by 2022, respectively (**Figure 4**). We observed a significant increase in the proportion of hospitalizations occurring at hospitals offering RPM services for both HF (OR of being hospitalized at RPM hospital, per year: 1.22; 95% CI: 1.22–1.23; p < .001) and AMI (OR, per year: 1.23, 95% CI: 1.23–1.24, p < .001).

**Figure 4.**
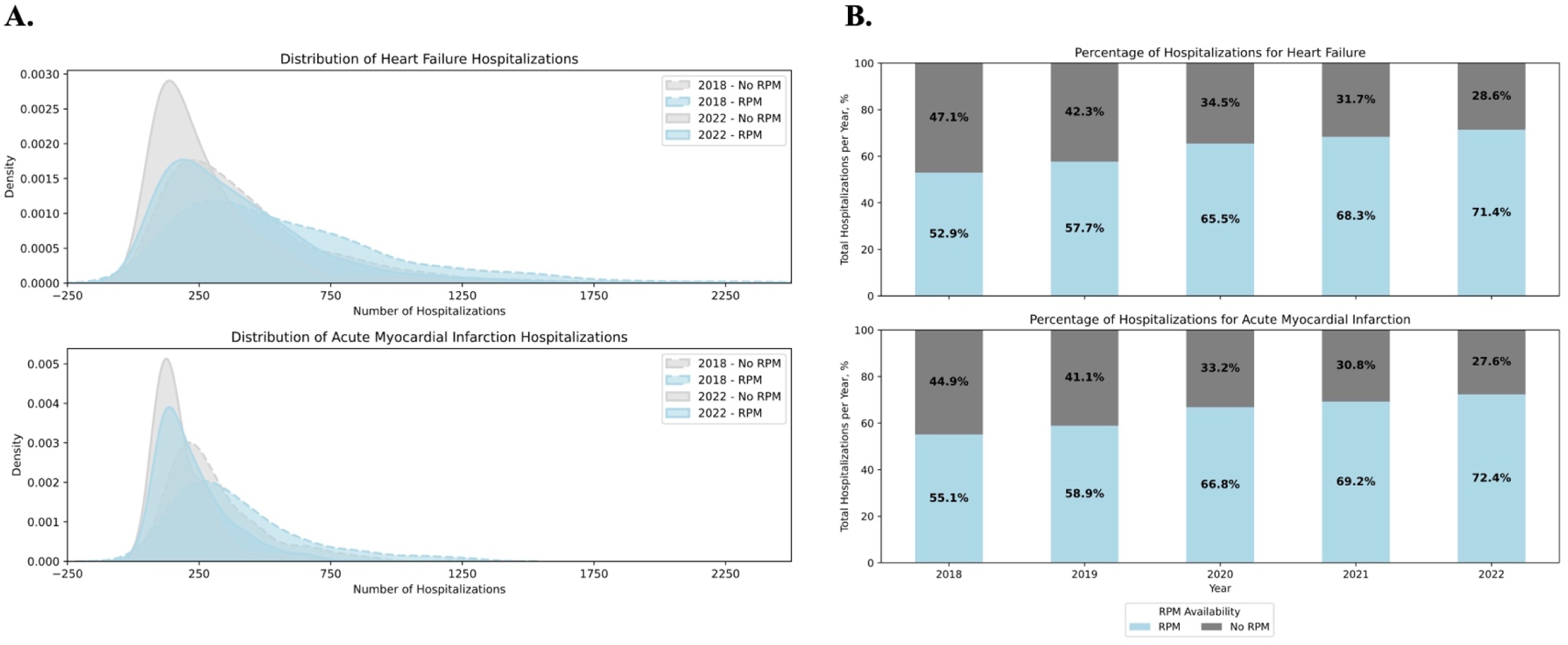
Trends in the Proportion of Hospitalizations for Heart Failure and Acute Myocardial Infarction at Hospitals with and without Remote Patient Monitoring (RPM), 2018–2022. **A.** Distribution of heart failure and acute myocardial infarction hospitalizations by RPM Status in 2018 vs 2022. **B.** Percentage of hospitalizations occurring in hospitals with and without RPM (2018–2022).

**Figure 5.**
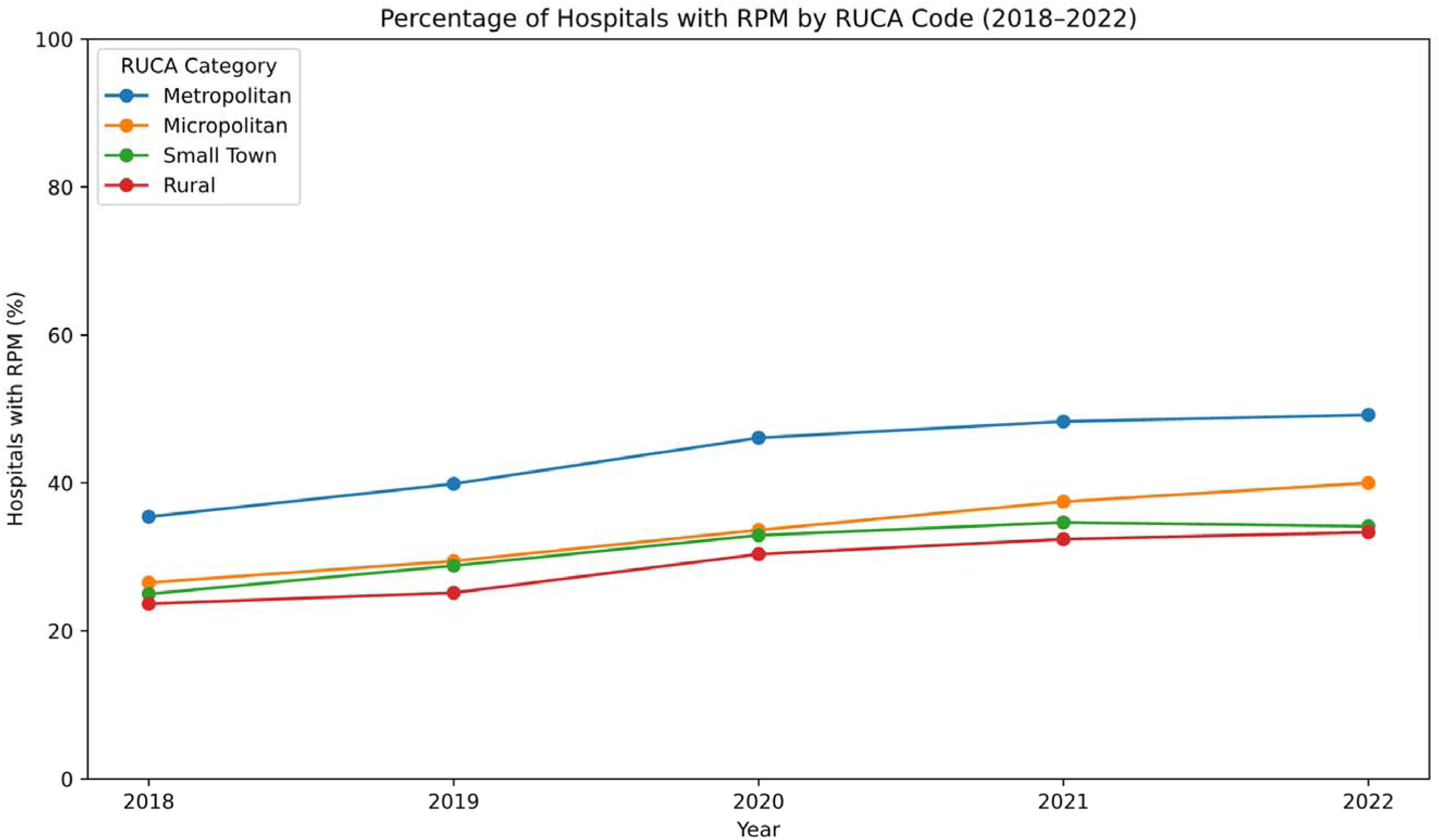
Trends in remote patient monitoring (RPM) service availability by rural-urban commuting area (RUCA) classification, 2018–2022.

## DISCUSSION

In this national study of US hospitals from 2018 through 2022, there was a 1.4-fold increase in the number of hospitals with RPM services, with nearly half of US hospitals offering these services by 2022. Notably, the majority of this growth occurred between 2018 and 2020 (a 30% relative increase), while the rate of expansion slowed significantly thereafter, with only a 7.9% relative increase from 2020 onward. This increase was observed for both post-discharge and chronic care RPM services. The availability of RPM services varied widely based on hospital and neighborhood characteristics, with lower availability of these services at small, rural, non-teaching hospitals, especially those in the South. Moreover, hospitals in communities with low household incomes and a large proportion of Hispanic individuals had substantially lower RPM availability. Across study years, we observed an increasing proportion of HF and AMI hospitalizations at hospitals offering RPM services, rising from approximately 50% in 2018 to over 70% in 2022, highlighting the expanding reach of RPM services.

Patient-level studies, like the one using insurance claims by Tang et al. (2022), have suggested a substantial recent increase in claims for RPM service in fee-for-service Medicare and Medicaid beneficiaries in the US between 2019 and 2021.^6,26^ This increase in the use of these services has occurred despite the lack of robust evidence on the clinical benefits of RPM or clarity on patients or conditions that benefit most from these services. Our findings complement the findings presented in these selected patient populations to provide evidence of the national landscape and describe the rapidly increasing proportion of US hospitals adopting RPM. However, we note that this uptake has varied widely across hospitals according to their size, type, and region of location. We observed that hospitals serving predominantly disadvantaged communities - that also consistently experience worse readmission outcomes^13^ – also have the lowest availability of RPM services. This national hospital-level analyses adds critical insight into the infrastructural capacities that enable or constrain the deployment of digital health technologies. The greater uptake of RPM services among larger, well-resourced hospitals may reflect their stronger institutional capacity to meet complex billing requirements, invest in digital infrastructure, and support the operational changes needed to scale remote monitoring, factors that may limit adoption in smaller or under-resourced settings and contribute to widening disparities in access.

These findings are pertinent given the growing evidence suggesting that emerging digital health technologies can improve patient outcomes.^18–21^ Numerous society guidelines emphasize the value of RPM services in enhancing appropriate post-discharge care.^3,22–24^ Specifically, many studies have suggested a role for guideline-directed therapy optimization in HF in the post-discharge period, while the critical services in AMI in this setting are less well-defined.^32,33^ Emerging randomized clinical trial evidence, however, suggests that RPM specifically designed to target post-acute coronary syndrome, including symptom-triggered monitoring on home wearable ECG, blood pressure, and pulse oximeter, with close coordination with a cardiologist, was associated with 76% lower readmission and emergency department visits over 6 months.^34^ These observations underscore the need to better characterize the nature of the services provided and their intended application to enable the evaluation of their impact on outcomes at hospitals.

This work also has important policy implications. The considerable investment in RPM services, further accelerated by the COVID-19 pandemic, has been supported by CMS and other payers through coverage in a fee-for-service model over the past five years.^12,35^ Besides the high availability and broad insurance coverage of RPM services, there is no standard definition for RPM and no consistent outcome testing.^12^ The observed heterogeneity in RPM service availability highlights that not all communities that could benefit from RPM have access to these services covered by national insurance providers. There is also a need to generate the necessary evidence for individual services to improve the efficiency of these investments. These considerations are especially critical as policymakers and payers consider the future of RPM coverage in the post-pandemic era.

Despite the introduction of new billing codes and financial incentives, RPM billing in the US remains highly concentrated among a small number of primary care providers.^28–30^ At the same time, health systems have faced considerable challenges in establishing financially sustainable, equitable, and scalable RPM programs.^9^ While current reimbursement models do not provide sufficient constraints on the nature of offered services, they include certain requirements, such as in-person device setup, minimum data transmission, and documentation of interactive communication, which can represent procedural constraints that may affect less resourced hospitals. Specifically, hospitals serving low-income populations, where patients may lack access to Bluetooth-enabled devices or may only be able to submit data manually, thereby failing to meet reimbursement criteria despite meaningful clinical participation. As illustrated by recent implementation studies,^31,32^ health systems have had to invest in device loaner programs and technical infrastructure to mitigate these limitations, yet still struggle to meet the criteria necessary for sustained billing under CPT codes. These payer-level constraints likely contribute to the geographic and structural disparities in RPM availability observed in our analysis.

Our study has certain limitations that merit consideration. First, we lack detailed information on the specific components of the RPM services evaluated. The RPM services reimbursed by payers under this category are not well-defined, underscoring the need to more specifically define the components of these services and their alignment with the available evidence for each condition. Second, while we have information on the characteristics of the communities served by these hospitals, we lack information on the number of patients and the nature of the conditions managed with these services at these hospitals. Effective implementation of RPM relies on patients having reliable internet access, suitable devices, digital literacy, and the capacity to engage with monitoring devices, factors often associated with higher socioeconomic status. Consequently, even within hospitals offering RPM, these services may predominantly benefit better-resourced patients who already have a lower baseline risk for adverse outcomes. Further investigations should focus on understanding the nature and quality of RPM services provided across different hospital types in the US, the specific patient populations benefiting from RPM services, the extent of their engagement, and the conditions most commonly managed using these technologies.

## CONCLUSION

In this national study of US hospitals, there has been a significant increase in the availability of RPM services over a recent 5-year period. However, there is a large variation in the availability of these services across hospitals and communities, with fewer hospitals serving low-income and rural communities offering these services. There is also an inconsistent association between RPM service availability at US hospitals and their readmission performance for cardiovascular conditions. These observations underscore the need to improve access to RPM services in vulnerable communities but with an emphasis on generating evidence essential for identifying the nature of services that enable the best patient outcomes.

## Data Availability

The data are available directly from the American Hospital Association and the Centers from Medicare and Medicaid Services.

https://github.com/CarDS-Yale/rpm_national

## Supplemental Materials

Supplemental Methods, Tables S1 and S2, Figures S1 to S4.

## Author Contributions

Drs Pedroso and Khera had full access to all of the data in the study and take responsibility for the integrity of the data and the accuracy of the data analysis. All authors approved the final version for submission. *Study concept and design*: Pedroso and Khera. *Acquisition, analysis, or interpretation of data*: Pedroso; Lin; Ross, and Khera. *Drafting of the manuscript*: Pedroso and Khera. *Critical revision of the manuscript for important intellectual content*: Lin; Ross, and Khera. *Statistical analysis*: Pedroso and Khera. *Obtained funding*: Khera. *Administrative, technical, or material support*: Pedroso and Khera. *Study supervision*: Khera.

