## Supplemental Methods for "National Patterns of Remote Patient Monitoring Service Availability at US Hospitals"

Rohan Khera MD, MS

#### Summary

**Supplementary Methods.** Data Sources.

**Table S1.** Association between hospital and community characteristics and availability of any remote patient monitoring (RPM) service or RPM targeted at post-discharge or chronic care.

**Table S2.** Hospitals without remote patient monitoring (RPM) services between 2018 and 2022. For individual hospital characteristics, the numbers and percentages represent those not offering RPM services in the respective hospital subgroups.

**Figure S1.** Relative increase in the availability of remote patient monitoring (RPM) services targeted at post-discharge care across hospital groups between 2018 and 2022.

**Figure S2.** Relative increase in the availability of remote patient monitoring (RPM) services targeted at chronic care across hospital groups between 2018 and 2022.

**Figure S3.** Community and hospital characteristics associated with the availability of remote patient monitoring (RPM) services for post-discharge care.

**Figure S4.** Community and hospital characteristics associated with the availability of remote patient monitoring (RPM) services for chronic care.

### Supplementary Methods

#### Data Sources

The American Hospital Association Annual Survey is a comprehensive data collection initiative administered to hospitals nationwide every year, with a primary objective to provide a database that supports research, policymaking, and benchmarking within the healthcare industry. The survey collects information directly from hospitals and healthcare systems, encompassing their organizational structure (e.g. type of organization responsible for establishing policy for overall operations and type of services provided to the majority of patients), care delivery architecture, and the availability of services.[14]

**Table S1.** Association between hospital and community characteristics and availability of any remote patient monitoring (RPM) service or RPM targeted at post-discharge or chronic care.

| | Any RPM | RPM, Post-discharge | RPM, Chronic care |
|---|---|---|---|
| | OR (95% CI) | | |
| Aged ≥65, % | 0.97 (0.87-1.08) | 0.92 (0.81-1.04) | 0.98 (0.88-1.10) |
| Female, % | 0.96 (0.87-1.05) | 1.01 (0.91-1.12) | 0.98 (0.90-1.08) |
| Black, % | 1.12 (1.00-1.26) | 1.16 (1.02-1.31) * | 1.08 (0.96-1.22) |
| Hispanic, % | 0.79 (0.68-0.93) * | 0.79 (0.66-0.94) * | 0.72 (0.60-0.85) * |
| Less than high school, % | 1.01 (0.88-1.16) | 1.06 (0.92-1.24) | 1.14 (0.99-1.32) |
| Median Household Income | 1.18 (1.05-1.32) * | 1.22 (1.08-1.37) * | 1.15 (1.02-1.29) * |
| Disabled, % | 0.91 (0.81-1.03) | 0.89 (0.78-1.02) | 0.85 (0.74-0.96) * |
| > 300 beds vs 100-300 | 2.07 (1.73-2.48) * | 2.13 (1.75-2.59) * | 1.83 (1.52-2.21) * |
| >300 beds vs <100 | 3.71 (2.90-4.74) * | 3.46 (2.69-4.45) * | 3.17 (2.49-4.05) * |
| Midwest vs Northeast | 0.79 (0.60-1.04) | 0.61 (0.47-0.80) * | 0.66 (0.50-0.86) * |
| South vs Northeast | 0.38 (0.29-0.49) * | 0.30 (0.23-0.40) * | 0.30 (0.23-0.39) * |
| West vs Northeast | 0.58 (0.43-0.78) * | 0.40 (0.29-0.55) * | 0.50 (0.37-0.68) * |
| Micropolitan vs Metropolitan | 1.21 (0.95-1.54) | 1.00 (0.76-1.31) | 1.23 (0.95-1.57) |
| Rural vs Metropolitan | 0.50 (0.32-0.77) * | 0.50 (0.32-0.77) * | 0.50 (0.32-0.77) * |
| Non-teaching vs Teaching | 0.29 (0.19-0.44) * | 0.34 (0.24-0.49) * | 0.30 (0.21-0.44) * |
| Private vs Government | 1.22 (0.97-1.54) | 1.62 (1.26-2.09) * | 1.29 (1.01-1.63) * |

*Statistically significant odds ratios (OR).

**Table S2.** Hospitals without remote patient monitoring (RPM) services between 2018 and 2022. For individual hospital characteristics, the numbers and percentages represent those not offering RPM services in the respective hospital subgroups.

| | Overall | 2018 | 2019 | 2020 | 2021 | 2022 |
|---|---|---|---|---|---|---|
| Hospitals, N | 5,644 | 4,129 | 3,948 | 3,865 | 3,853 | 3,883 |
| Hospitals with RPM services | 3,446 (61.1%) | 2,765 (67.0%) | 2,481 (62.8%) | 2,207 (57.1%) | 2,106 (54.7%) | 2,086 (53.7%) |
| **Bed size** | | | | | | |
| <100 | 2247 (70.8%) | 1646 (75.9%) | 1501 (72.7%) | 1395 (68.6%) | 1335 (66.1%) | 1342 (65.8%) |
| 100-300 | 932 (56.9%) | 784 (64.1%) | 692 (59.1%) | 583 (51.6%) | 556 (49.2%) | 548 (47.9%) |
| >300 | 267 (32.1%) | 335 (45.5%) | 288 (40.3%) | 229 (32.6%) | 215 (30.6%) | 196 (28.0%) |
| **US Region** | | | | | | |
| Northeast | 365 (51.0%) | 278 (52.1%) | 220 (44.6%) | 194 (39.4%) | 184 (37.6%) | 179 (37.2%) |
| Midwest | 843 (53.9%) | 766 (62.0%) | 692 (58.5%) | 608 (51.4%) | 599 (51.3%) | 557 (48.9%) |
| South | 1518 (68.6%) | 1227 (75.1%) | 1128 (71.8%) | 1071 (68.0%) | 988 (64.4%) | 985 (63.8%) |
| West | 661 (60.8%) | 490 (67.8%) | 438 (62.6%) | 323 (53.5%) | 319 (49.6%) | 351 (50.0%) |
| **Area Type** | | | | | | |
| Metropolitan | 2203 (58.6%) | 1727 (63.1%) | 1536 (58.7%) | 1321 (51.9%) | 1283 (49.8%) | 1275 (49.1%) |
| Micropolitan | 519 (63.3%) | 443 (71.7%) | 416 (68.4%) | 388 (64.1%) | 364 (61.9%) | 341 (59.0%) |
| Rural | 724 (67.9%) | 595 (76.7%) | 529 (73.0%) | 498 (69.7%) | 459 (66.5%) | 470 (66.6%) |
| **Teaching** | | | | | | |
| Yes | 33 (12.8%) | 80 (32.0%) | 66 (26.9%) | 41 (16.9%) | 36 (15.0%) | 33 (13.7%) |
| No | 3413 (63.4%) | 2685 (69.2%) | 2415 (65.2%) | 2166 (59.8%) | 2070 (57.3%) | 2053 (56.4%) |
| **Ownership** | | | | | | |
| Government | 775 (67.7%) | 609 (76.9%) | 541 (72.6%) | 489 (67.2%) | 456 (62.9%) | 435 (61.1%) |
| Private | 2671 (59.4%) | 2156 (64.6%) | 1940 (60.6%) | 1718 (54.8%) | 1650 (52.7%) | 1651 (52.1%) |

**Figure S1.** Relative increase in remote patient monitoring (RPM) services availability targeted at post-discharge care across hospital group between 2018 and 2022.

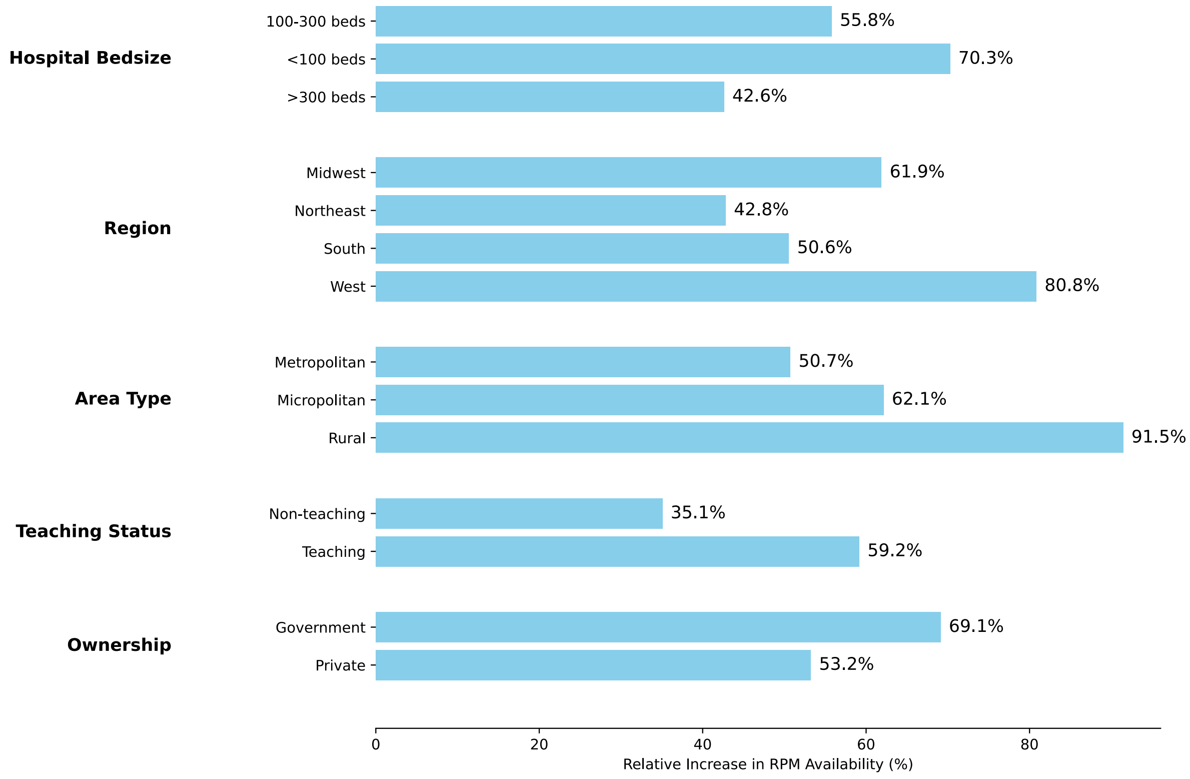

**Figure S2.** Relative increase in remote patient monitoring (RPM) services availability targeted at chronic care across hospital group between 2018 and 2022.

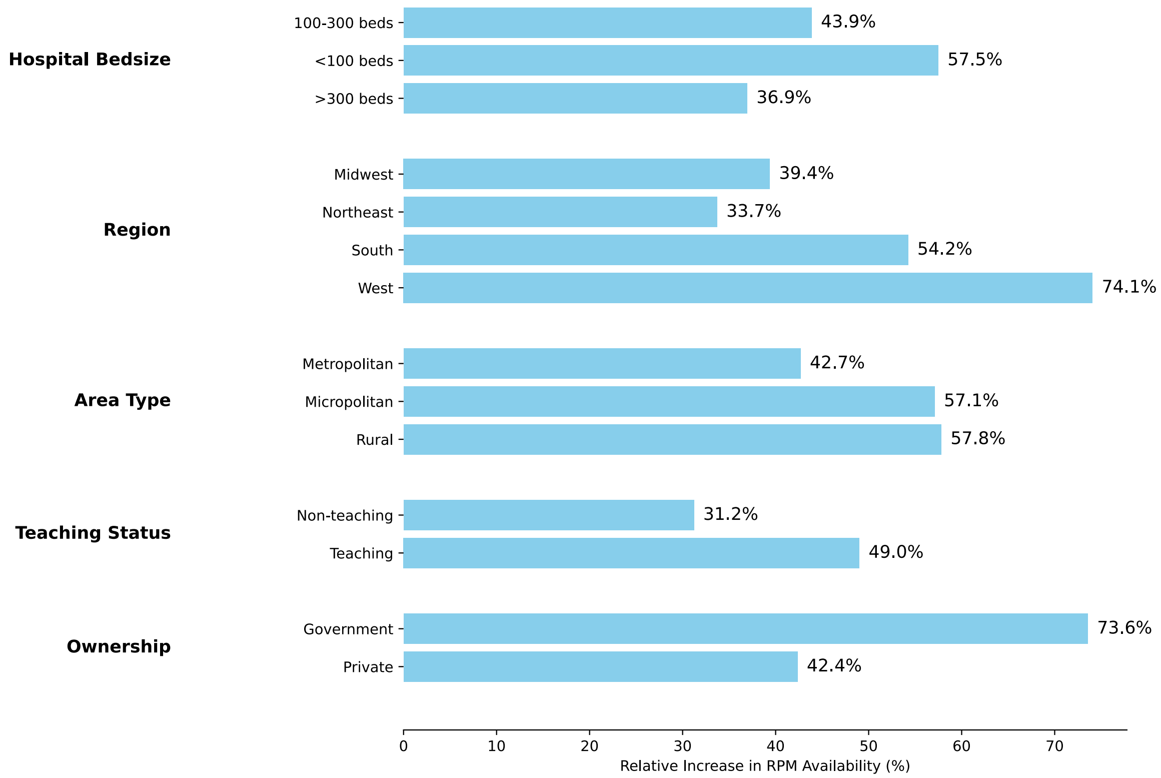

**Figure S3.** Community and hospital characteristics associated with the availability of remote patient monitoring (RPM) services for post-discharge care.

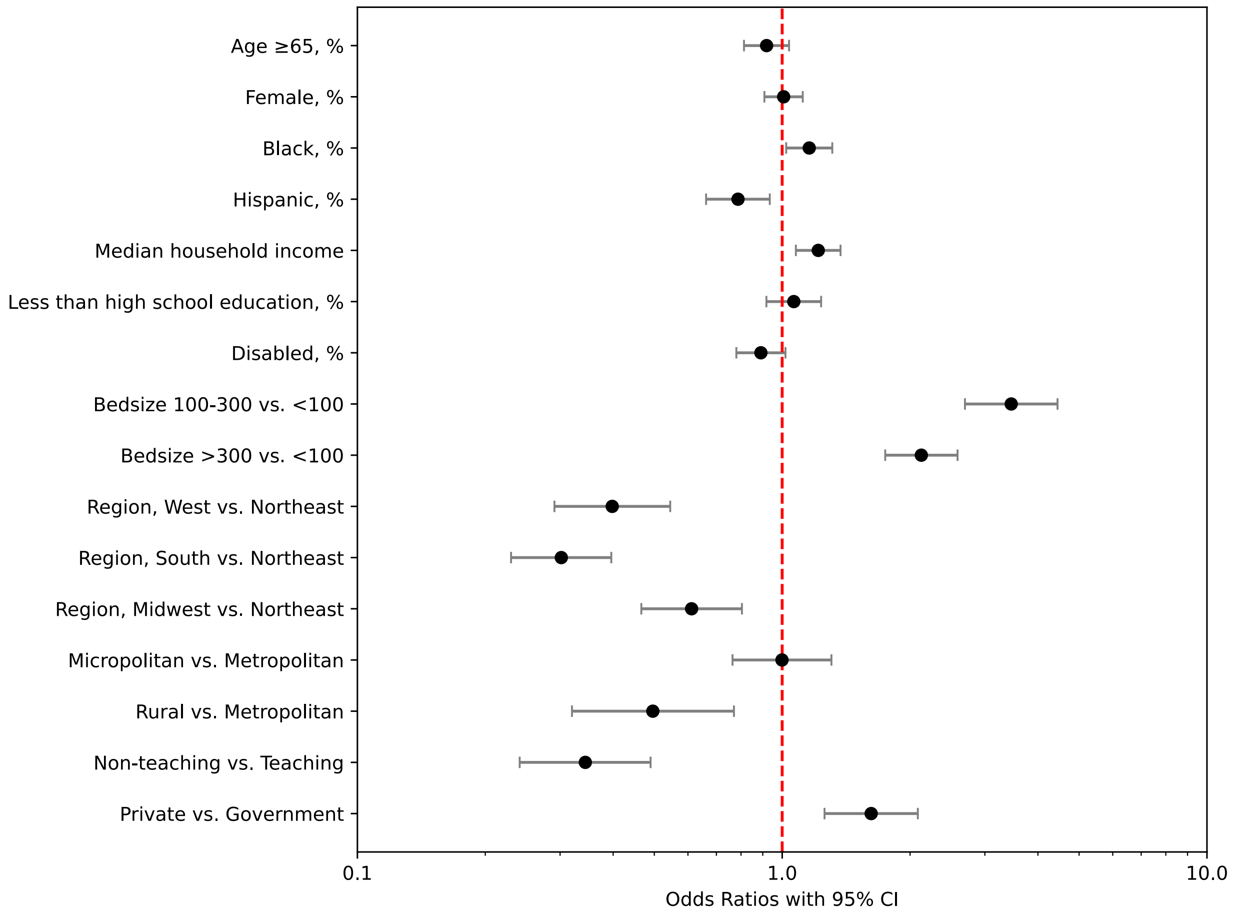

**Figure S4.** Community and hospital characteristics associated with the availability of remote patient monitoring (RPM) services for chronic care.

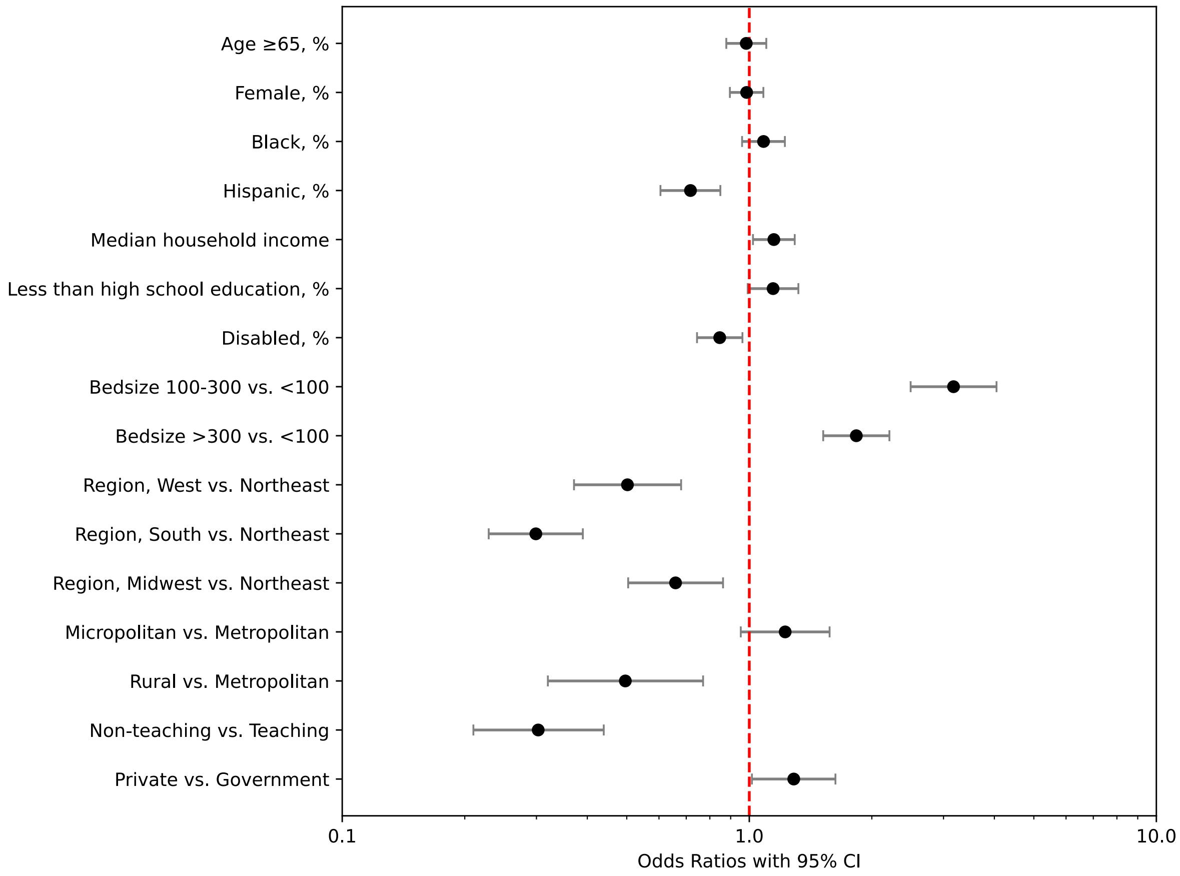
